# Thalamocortical network neuromodulation for epilepsy

**DOI:** 10.1101/2024.02.12.24302647

**Authors:** Shruti Agashe, Juan Luis Alcala-Zermeno, Gamaleldin M. Osman, Keith Starnes, Benjamin H. Brinkmann, Doug Sheffield, Kent Leyde, Matt Stead, Kai J. Miller, Jamie J. Van Gompel, Gregory A. Worrell, Brian N. Lundstrom, Nicholas M. Gregg

**Affiliations:** Department of Neurology, Mayo Clinic, Rochester, MN, USA; Department of Neurology, Duke University, NC, USA; Department of Neurology, Thomas Jefferson University, Philadelphia, PA, USA; Division of Child Neurology, Department of Pediatrics, McGovern Medical School at UTHealth, Houston, TX, USA; Cadence Neuroscience, Redmond, WA, USA; Dark Horse Neuro, Inc., Bozeman, MT, USA; Department of Neurosurgery, Mayo Clinic, Rochester, MN, USA

**Keywords:** Neuromodulation, deep brain stimulation, cortical stimulation, anterior nucleus of the thalamus, centromedian nucleus of the thalamus

## Abstract

**Objectives:** Despite the growing interest in network-guided neuromodulation for epilepsy, uncertainty about the safety and long-term efficacy of thalamocortical stimulation persist. Our evaluation focused on the use of a 4-lead open-loop implantable pulse generator (IPG) for thalamocortical network neuromodulation.

**Methods:** We retrospectively reviewed seven subjects with diverse seizure networks (SNs)—poorly localized, regional, or multifocal—undergoing thalamocortical neuromodulation. Employing a 4-lead system, electrodes targeted both thalamic and cortical SN nodes. We assessed seizure severity, life satisfaction, and sleep quality on a 10-point scale, and seizure frequency via telephone interviews and chart review. Six subjects underwent open-loop stimulation trials during intracranial EEG (iEEG) to confirm SN engagement and optimize settings, targeting the suppression of interictal epileptiform discharges (IEDs) and seizures. Outcomes were assessed by Wilcoxon sign-rank test, 0.05 significance level.

**Results:** After a median of 17 months post-implantation (range 13–60), subjects had a median disabling seizure reduction of 93% (range 50-100%, p=0.0156), with 100% responder rate (≥50% reduction in seizure frequency). The median improvement in seizure severity was 3.5 out of 10 points (p=0.0312), life satisfaction 4.5 points (p= 0.0312), and quality of sleep 3 points (p=0.062). No perioperative complications occurred. Rare transient seizure exacerbations and stimulation-related sensory/motor side effects resolved with parameter adjustments. One subject required surgical revision due to delayed infection. Six subjects had permanent electrode placement refined by iEEG trial stimulation; five subjects had >90% reduction in seizure frequency during iEEG stimulation.

**Significance:** Thalamocortical network neuromodulation using a 4-lead open-loop system is safe, and is associated with significant improvements in seizure control and patient quality of life. Trial stimulation during iEEG shows promise for enhancing SN engagement and parameter optimization, but requires further study. Prospective controlled trials are needed to establish the validity of thalamocortical network neuromodulation for epilepsy.

**Key Points:**

- Thalamocortical neuromodulation with a 4-lead open-loop stimulation system is feasible and safe, and is associated with significant improvements in seizure control and life satisfaction.
- Trials of therapeutic stimulation during phase 2 iEEG monitoring has the potential to refine seizure network engagement and optimize stimulation parameters, for more effective chronic neuromodulation.
- Prospective controlled trials are needed to validate the efficacy of thalamocortical network neuromodulation.

## Introduction

Drug resistant epilepsy (DRE) affects approximately one third [1, 2] of individuals with epilepsy. For individuals with DRE focal epilepsy who are poor surgical candidates due to diffuse seizure networks (SN), multiple foci, or overlap with eloquent cortex, neuromodulation is a viable treatment option. There are multiple intracranial neuromodulation paradigms that have been used for epilepsy in clinical practice, including FDA-approved responsive neurostimulation (RNS)[3], anterior thalamic nuclei deep brain stimulation (ANT-DBS) [4-7], and the off-label use of chronic subthreshold stimulation (CSS) ([8-11]), with additional novel approaches supported by preclinical work[12]. In addition to differences in stimulation targets between DBS for epilepsy (subcortical structures, typically thalamus) and RNS (typically cortex), there are differences in the stimulation paradigms. Open-loop stimulation (DBS and CSS) delivers therapy at a set frequency and schedule regardless of changing brain activity, while closed-loop stimulation (RNS) delivers therapy in response to detected changes in the electrocorticography signal. Open-loop therapy can be delivered as continuous electrical stimulation, e.g. 2 Hz, or as a duty cycle therapy, e.g. 145 Hz with cycling of 1 minute on and 5 minutes off. Previous investigations have shown that open-loop cortical CSS stimulation using a 4-lead IPG, like the approach used here, is effective in reducing interictal epileptiform discharges (IEDs) and seizures [21].

There is a growing body of evidence that indicates epilepsy is a disorder of brain networks[13, 14], and that cortico-thalamo-cortical circuits are involved in the onset, maintenance, and propagation of seizures [13, 15-21]. This progress has led to great interest in network guided neuromodulation for epilepsy[22], however, little is known about combined cortical and thalamic SN node stimulation. Recent work with the RNS system suggests there is potential utility in combined cortical plus thalamic responsive neurostimulation to reduce seizure frequency. In a three subject series by Elder et al., a single cortical strip combined with unilateral ANT stimulation resulted in a ≥50% reduction in seizure burden in 2 subjects, and was overall found to be safe and effective[23]. Two series by Burdette et al., [24, 25] of cortical plus centromedian nucleus, and cortical plus medial pulvinar nucleus stimulation, reported ≥50% seizure reduction. The thalamic centromedian nucleus (CM) [26-28] and medial pulvinar[29] are DBS targets of interest for the treatment of extra limbic and generalized epilepsy, and posterior quadrant and neocortical temporal lobe epilepsy, respectively.

In addition to growing interest in network guided neuromodulation, efforts are underway to characterize the short latency effects of electrical stimulation on seizure networks and brain excitability [10, 30-32] to inform neuromodulation, here termed Biomarker Targeted Stimulation (BTS). As defined here, BTS consists of two phases, comprised of 1) a trial of electrical brain stimulation delivered to the SN via intracranial EEG (iEEG) monitoring electrodes, to refine SN targets and stimulation paradigms, followed by 2) chronic device implantation and treatment, guided by phase 1 trial stimulation. During BTS phase 1, stimulation targets, paradigms, and parameters are guided by objective modifiable biomarkers, (such as the suppression of interictal biomarkers (i.e. IEDs) and seizures). BTS trial stimulation could include responsive, duty-cycle, continuous, Poisson distributed, and other novel stimulation paradigms, delivered to various cortical, subcortical grey or white matter targets.

In this retrospective case-series we evaluate the safety, feasibility, and efficacy of thalamocortical network neuromodulation using an open-loop 4-lead system, in a cohort of individuals with poorly localized, multifocal (more than 2), eloquent cortex, or regional SNs. Chronic open-loop stimulation was delivered using FDA approved 4-lead IPGs in an off-label manner, similar to prior work (published as CSS) [8-11]. Additionally, we describe the results of BTS trial stimulation in this cohort, including safety, feasibility, and impact on SN biomarkers.

## Methods

This is a retrospective case series study. This series reports the first 7 subjects at our institution treated with open-loop 4-lead thalamocortical network neuromodulation. Group averaged partial long-term outcome data from 6 of these subjects were published in prior work comparing CSS (n=32 subjects, 28 with cortex only leads), ANT-DBS, CM-DBS, RNS, and VNS [33]. This work extends beyond the prior publication, reporting subject specific data and analysis of long-term outcome data for thalamocortical network neuromodulation, individualized characterization of cortical and thalamic targeting and stimulation parameters, and quantitative analysis of BTS trial stimulation modulation of biomarkers during phase 2 iEEG. This retrospective study was approved by the Mayo Clinic Institutional Review Board. Cohort characteristics are described in Table 1. All subjects had a comprehensive epilepsy evaluation that included video EEG monitoring, high resolution MRI, and iEEG monitoring. Subjects had focal DRE according to ILAE criteria [2] with SN not amenable to surgical resection due to overlap with eloquent cortex, or SN that were poorly localized, regional, or multifocal (2 or more foci) based on iEEG.

**Table 1.** Baseline characteristics, imaging findings, seizure networks, final implant locations and duration of follow-up for study cohort. M:Male, F: Female, L:Left, R: Right, SO: Seizure onset, ASMs: Antiseizure medications, FIAS: focal impaired awareness seizure, FAS: Focal awareness seizure, FBTCS: Focal to bilateral tonic clonic seizure, ANT: Anterior nucleus of the Thalamus, CM: Centromedian nucleus of the thalamus VIM: Ventral intermediate nucleus of thalamus, VNS: vagus nerve stimulator.

| Subject | Age at implant | Sex | Age at onset (yrs) | Semiology | ASMs tried | ASMs/devices at implant | MRI/PET | Seizure Network | Localization | Permanent lead locations | Follow-up duration |
| --- | --- | --- | --- | --- | --- | --- | --- | --- | --- | --- | --- |
| 1 | 26-30 | M | 16-20 | FAS motor, FBTCS | 2 | Levetiracetam, Lamotrigine | MRI: right frontal, temporal and parietal encephalomalacia secondary to perinatal stroke | Right frontal operculum, insula | Poorly localized, regional | R ANT + anterior insula + 2 right frontal operculum | 60 months |
| 2 | 21-25 | F | 6-10 | FAS motor, FIAS non-motor, FBTCS | 13 | Oxcarbazepine | MRI: non lesional, PET: Left mesial frontal hypometabolism | Left middle frontal, left cingulate, left precentral gyrus | Eloquent cortex, regional | L ANT + L superior frontal + L mid cingulate + L medial frontal gyrus | 24 months |
| 3 | 11-15 | M | 0-5 | FAS motor, FIAS non-motor | 3 | Clobazam, Lacosamide, Oxcarbazepine | MRI: non lesional, PET: non lesional | Left anterior and mid-cingulate, left mesial frontal, left posterior insula | Multifocal, regional | L ANT + L anterior insula + L posterior insula + mid cingulate | 13 months |
| 4 | 21-25 | F | 6-10 | FAS non-motor, FBTCS | 6 | Clonazepam, Lacosamide, Topiramate | MRI: non lesional, PET: non lesional | Right frontal and parietal | Eloquent cortex, regional | R ANT + R precentral + R postcentral + R paracentral lobule | 17 months |
| 5 | 41-45 | M | 16-20 | FIAS motor, FBTCS | 1 | Levetiracetam, Lamotrigine, Lacosamide | MRI: left temporal and frontal lobes possible focal cortical dysplasia, PET: non lesional | Left posterior temporal | Poorly localized, regional | L ANT + L superior temporal gyrus + L middle temporal gyrus + L inferior temporal gyrus | 24 months |
| 6 | 16-20 | F | 6-10 | FIAS motor | 7 | Cenobamate, Clonazepam | MRI: non lesional, PET: non lesional | Right frontal and parietal | Eloquent cortex | R CM + R precentral + R mesial precentral + R post central | 8 months |
| 7 | 30 | M | 0-5 | FIAS motor, FBTCS | 8 | Oxcarbazepine, Brivacetam, VNS | MRI: non lesional | Right mesial frontal and perirolandic | Eloquent cortex, regional | R ANT + R VIM + R mesial frontal + R perirolandic | 16 months |

All subjects had intracranial monitoring with stereo EEG (sEEG) except subject 5 who had subdural grid and depth electrodes monitoring. Subject 7 independently underwent both sEEG and grid implantation.

All cases were discussed at our multidisciplinary epilepsy surgery conference including epilepsy, neurosurgery, neuroradiology, and neuropsychology staff. FDA-approved systems and off-label neuromodulation 4tereotact were discussed with all patients. These individuals with poorly localized, regional, multifocal, and eloquent cortex SNs proceeded with off-label 4-lead open-loop thalamocortical network neuromodulation. Cortical electrodes targets were informed by phase 2 iEEG BTS trial stimulation; thalamus targets were primarily based on normative thalamocortical connectivity (ANT for frontotemporal networks, CM for peri-rolandic networks).

A presurgical localizing stereotactic MRI was performed followed by 5tereotacticc placement of chronic electrodes using a rigid Leksell frame. Thalamic targeting used a combination of direct and indirect methods, including direct visualization of the ANT using white matter nulled MRI sequence (FGATIR) [35], anterior commissure-posterior commissure (AC-PC) based offsets, and Montreal Neurological Institute (MNI) template space Krauth/Morel atlas structures[36] warped into patient space. Details regarding targeting of CM nucleus have been discussed in our prior work [26, 37].

The chronic implant used FDA approved neuromodulation hardware in an off-label manner (IPG: Boston Scientific Vercise Genus DBS R32 (4 leads, 32 electrode contacts) [38] or Medtronic Intellis (4 leads, 16 electrode contacts); Electrodes: Boston Scientific DB-2201-45 and DB-2202-45, Medtronic 3387, Medtronic 3391). Six subjects had 3 cortical leads and one thalamic lead. Subject 1-5 had one ANT lead, subject 6 (peri-rolandic SN) had a CM lead. Subject 7 with a medial frontal and peri-rolandic SN had 2 thalamic (ANT and ventral intermediate nucleus (Vim; projects to primary motor cortex)) and 2 cortical leads (Table 2, Figure 1).

**Table 2:**
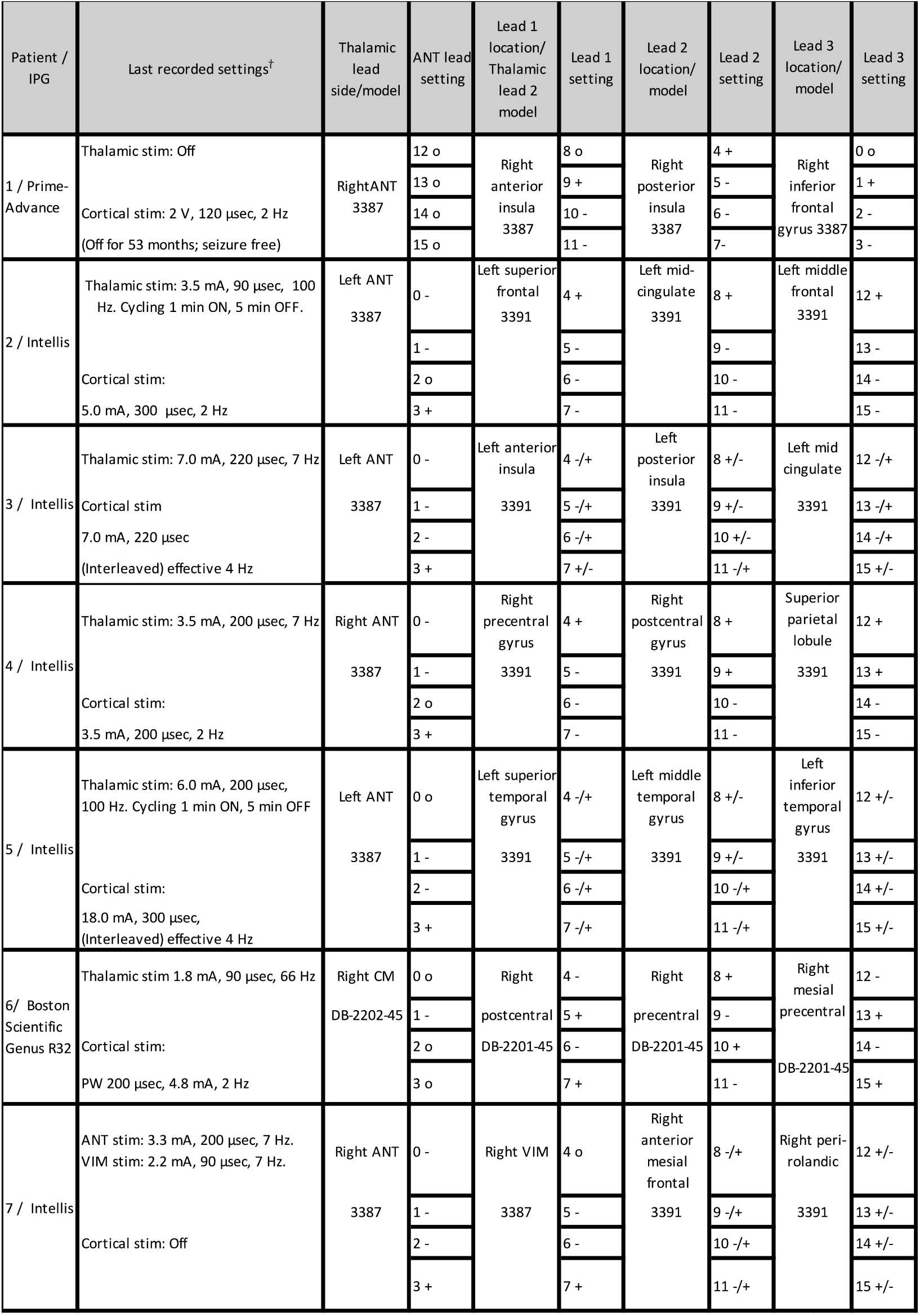
Stimulation programs at last follow up. Interleaved settings are represented as +/- or -/+ on each lead. Contacts 0, 4, 8, 12 are distal. For Subject 6, every other contact was numbered and programed for cortical stimulation; thalamic stimulation was referential with pulse generator serving as anode. Continuous stimulation was used unless noted otherwise. ANT=anterior nucleus of the thalamus. CM=centromedian nucleus. IPG=Implantable Pulse Generator, VIM=ventral intermediate nucleus.

**Figure 1:**
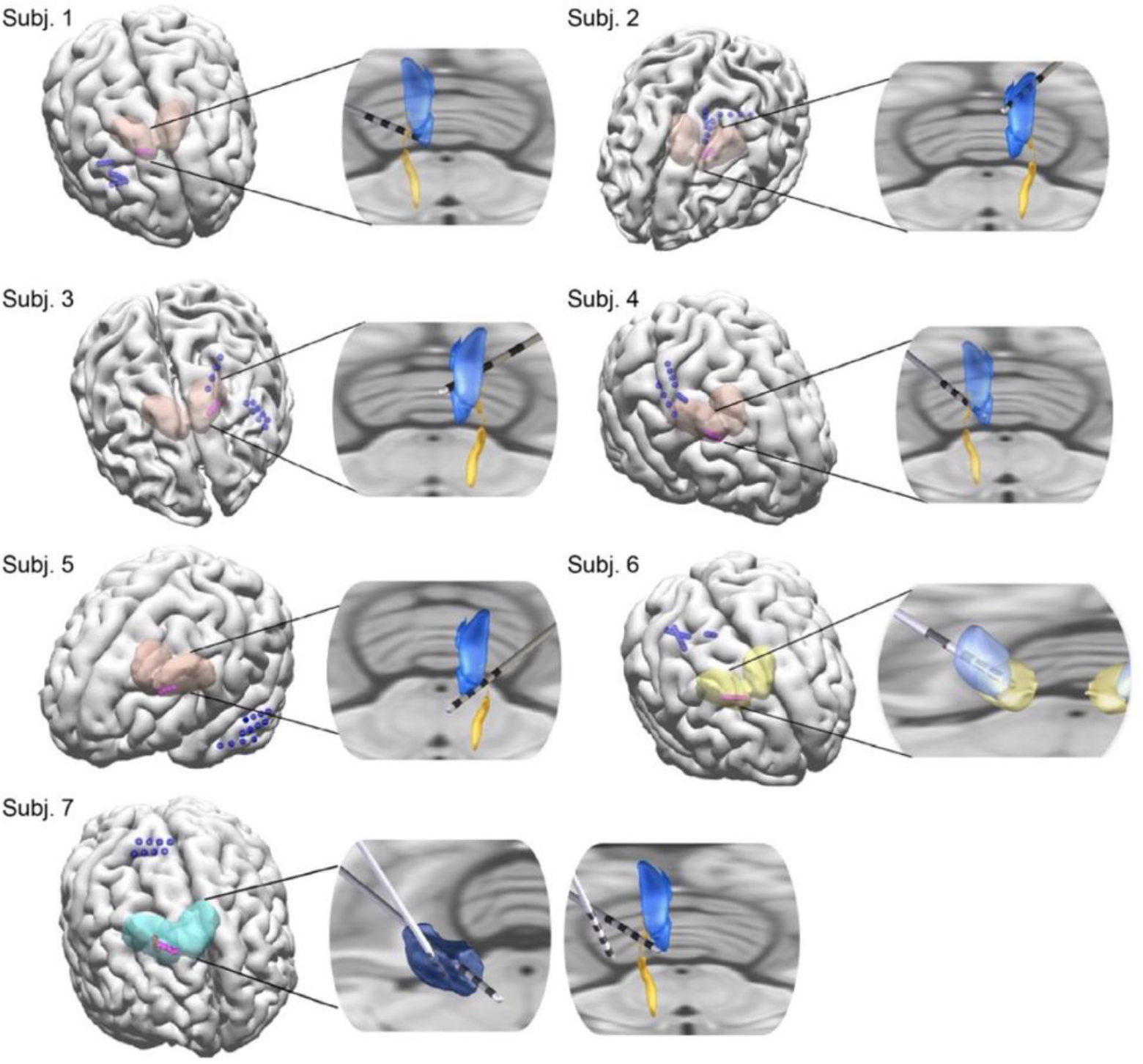
3D rendering (pink-thalamic leads, blue-cortical leads) with inset axial sections with rendered thalamic leads and nuclei (ANT (blue), mammillothalamic tract (yellow), subject 6: CM (light blue)-parafascicular nucleus (gold), and subject 7: VIM (dark blue)), using Curry and Lead-DBS imaging packages.

Postoperative head CT coregistered to preoperative MRI demonstrated well positioned leads using the Lead-DBS imaging package [39] (https://www.lead-dbs.org) and Krauth/Morel atlas structures demonstrated well positioned thalamic leads [26, 37] (Figure 1).

A BTS stimulation trial was initiated at the end of intracranial monitoring (or prior to lead internalization for subject 1) in six of seven subjects to aid in cortical stimulation target selection and parameter adjustment. Subject 7 received chronic 4-lead thalamocortical network neuromodulation and so is included in this case series, but did not complete a BTS stimulation trial. Stimulation was delivered through the most active intracranial electrodes identified to be in the SN, using an open-loop external neurostimulator module (Medtronic 37022), targeting up to 16 electrode contacts. Stimulation parameters ranged between 120-300 μsec pulse width, 2 Hz, and 0.3-3 V (typical bipolar impedance 1-2.5 kΩ; stimulation montage was most often 8 stimulated bipolar pairs), with stimulation trials lasting between 21 and 51 hours. Stimulation contacts and parameters adjustments were made to refine stimulated contact selection and parameters, based on short latency biomarkers (IEDs) and seizures (i.e. biomarker targeted stimulation) (Figure 3).

Clinical decision-making was based on qualitative assessment of IEDs and seizure counts. Here, we report results from an automated IED detector [34] to quantitatively assess the change in IED frequency in wake/sleep state-matched epochs during baseline and trial stimulation periods. For subject 1, quantification of the interictal burden was based on visual analysis due to ubiquitous stimulation artifacts confounding the automated detector (limited pre-internalization stimulation trial through the chronic system; no neighboring non-stimulated pairs to analyze). Baseline seizure counts are the number of seizures that occurred in the 24 hours preceding initiation of the BTS stimulation trial; BTS stimulation trial seizure rate was the number of seizures that occurred over 24 hours (21 hours for subject 1) at optimized stimulation parameters. BTS stimulation trials prioritized 1) optimization of cortical stimulation targets, followed by 2) parameter titration.

Seizure frequency data were recorded from chart review at the last follow-up. The seizure severity, life satisfaction, and quality of sleep scores were assessed by a 10 point scale through telephone interview, as prior [33] (subjects 1-5 and 7 listed as CSS in that publication) (Table 3, Figure 2). Response was defined as a ≥50% reduction in seizure rate. Statistical significance was assessed using the two-sided Wilcoxon signed rank test, with 0.05 significance level. Analyses were performed using MATLAB, version 2020b, Mathworks Inc, Natick, MA, USA.

**Table 3:** Chronic thalamocortical network neuromodulation outcomes, reporting monthly seizure frequency at pre-implantation baseline and at last follow-up for all subjects. Seizure severity, life satisfaction, and quality of sleep scores are listed (not available for subject 6).

| Patient | Follow-up (mo) | Disabling seizures/mo. |  |  | Seizure severity (worst = 10) |  |  | Life satisfaction (best = 10) |  |  | Quality of sleep (best = 10) |  |  |
| --- | --- | --- | --- | --- | --- | --- | --- | --- | --- | --- | --- | --- | --- |
|  |  | Before | After | % Improve | Before | After | Change | Before | After | Change | Before | After | Change |
| 1 | 60 | 120 | 0 | 100 | 8 | 1 | 7 | 3 | 10 | 7 | 5 | 5 | 0 |
| 2 | 24 | 14.5 | 0.75 | 95 | 5 | 3 | 2 | 7 | 9 | 2 | 7 | 10 | 3 |
| 3 | 13 | 135 | 10 | 93 | 7 | 4 | 3 | 6 | 4 | 2 | 3 | 5 | 2 |
| 4 | 17 | 7 | 2 | 71 | 10 | 8 | 2 | 3 | 7 | 4 | 2 | 8 | 6 |
| 5 | 24 | 210 | 100 | 52 | 8 | 4 | 4 | 9 | 3 | 6 | 4 | 7 | 3 |
| 6 | 16 | 7 | 0.5 | 93 | NA | NA | NA | NA | NA | NA | NA | NA | NA |
| 7 | 16 | 90 | 45 | 50 | 6 | 2 | 4 | 3 | 8 | 5 | 4 | 7 | 3 |

**Figure 2:**
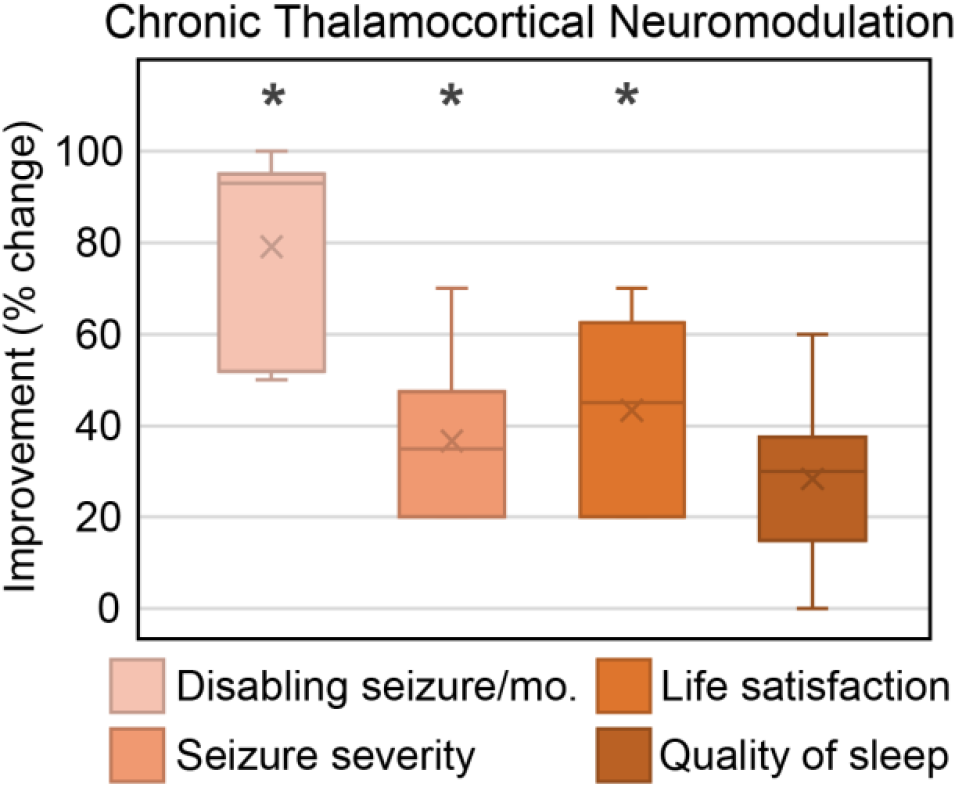
Box whisker plots of Chronic thalamocortical network neuromodulation outcomes; mean marked by ‘x’ and median by horizontal line. Change in seizure severity, life satisfaction, and quality of sleep adjusted to 100 point scale. Statistically significant results indicated by ‘*’.

## Results

Seven subjects with drug-resistant epilepsy, median age at implant 22 (range 14-42), received chronic open-loop thalamocortical network neuromodulation. The median follow-up time was 17 months (range 13-60 mo.).

The permanently implanted leads are shown in Figure 1. Electrode targets and stimulation parameters at last follow up are in Table 2. Following system placement, subject 1 received stimulation for 7 months; he experienced seizure reduction in the first 5 months followed by 2 months of seizure freedom while on stable antiseizure medication regimen. Stimulation was then inadvertently turned off, with sustained seizure freedom for the next 53 months through last follow-up, which we reported previously [40]. Subject 7 reported an initial increase in seizure frequency and was transitioned to thalamus only stimulation.

Chronic thalamocortical neuromodulation was associated with a significant reduction in seizure frequency and severity, with 93% median reduction in disabling seizures (range 50-100, p = 0.0156), and 3.5 point median improvement in seizure severity score (range 2-7, p=0.0312) (Table 3, Figure 2). There was 100% responder rate. Life satisfaction improved by 4.5 points (range 2-7, p=0.0312). Quality of sleep had nonsignificant change, with median improvement of 3 points (range 0-6, p=0.0625). Seizure severity, sleep quality, and life satisfaction were assessed by a standardized survey at a single timepoint, not necessarily at last follow-up.

Six subjects had a BTS stimulation trial through iEEG monitoring electrodes prior to chronic system implantation, or through externalized chronically implanted leads prior to system internalization (subject 1). High channel count iEEG (up to 256 channels) during trial stimulation allowed for stimulation target and parameter optimization to guide the chronic implant therapy.

Median baseline seizure burden in the 24 hours prior to initiation of trial stimulation was 14 (range 2-44). The median reduction in IEDs was 29% (range 0-87%, p = 0.062) and the median seizure reduction was 100% (range 50-100%, p = 0.031) at optimized settings. See supplemental information for trial stimulation details, including stimulation contact configurations, parameter adjustments, and antiseizure medications. Subjects 3, 5, and 6 were on stable antiseizure medications throughout baseline and trial stimulation periods; subjects 1 and 4 received intravenous lorazepam (clustered seizures) and intravenous lacosamide (preceding stimulation functional mapping), respectively, between the baseline and trial stimulation period. Subject 2 was on oxcarbazepine 600 mg twice daily at baseline, and 900 mg twice daily during trial stimulation. Figure 3 shows a sample 25 second epoch from subject 3 at baseline and after biomarker targeted stimulation trial. The trial stimulation technique has been discussed previously [8-11]. Cortical stimulation functional mapping was performed in 5 subjects.

**Figure 3.**
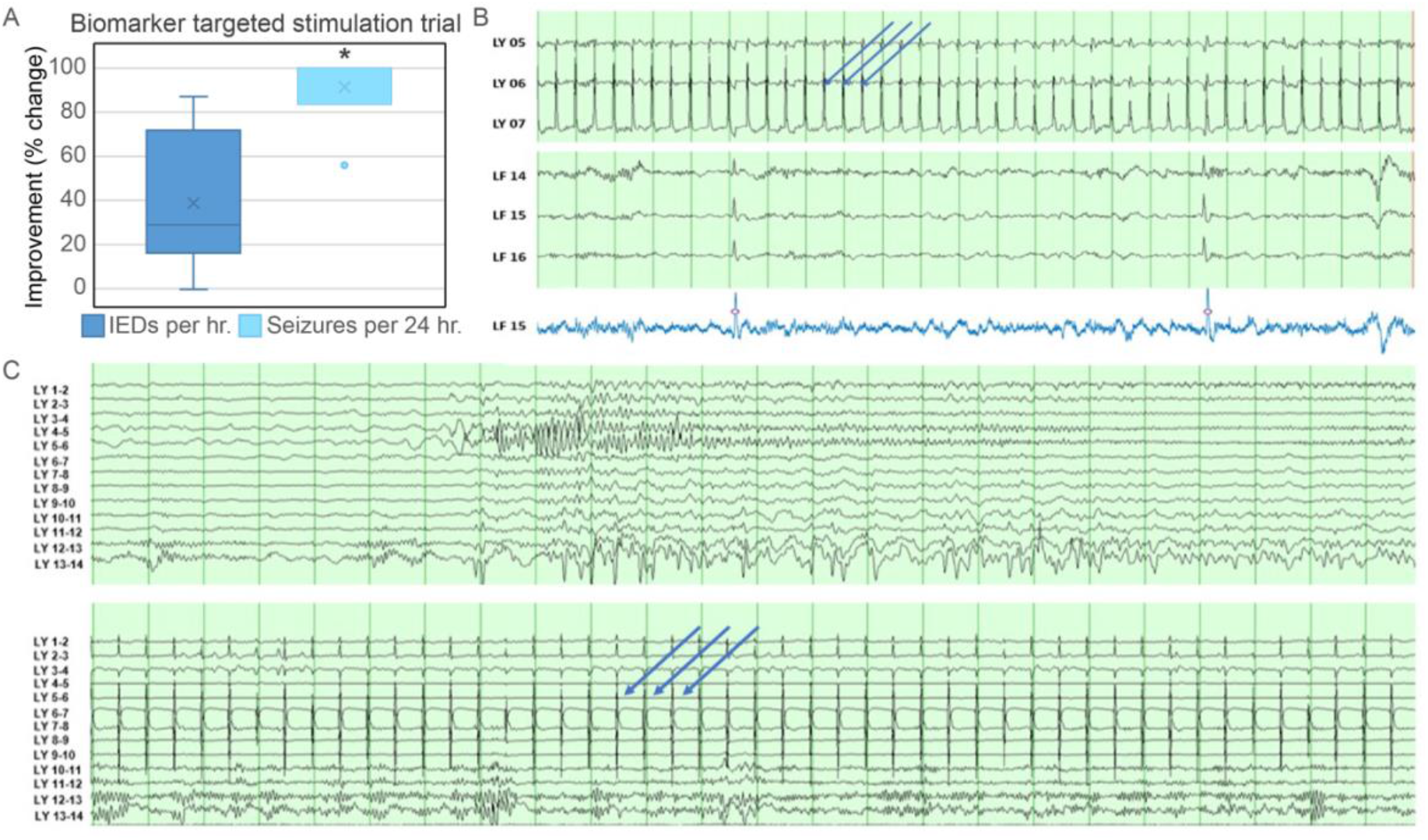
A) Box and whisker plot showing results from trial stimulation for 6 subjects; statistically significant results indicated by *. B) Top two panels showing intracranial EEG with 2Hz stimulation artifact (blue arrows) in LY (medial anterior cingulum) and IEDs in LF (inferior frontal/gyrus rectus); the bottom panel shows output from automated IED detector [23] on a single channel (LF 15, blue tracing; third-order one-dimensional median filtering to remove stimulation artifact). Automated IED detections marked with ‘o’. C) Sample intracranial recording during sleep from subject 3, bipolar montage, top panel (pre-stimulation, showing seizure discharge from LY 4-5 and 6-7 causing frequent arousals) and bottom panel matching lead and sleep state post stimulation at 2 Hz (blue arrows stimulation artifact), 2 V, 200 μsec. time base 15 mm/sec, sensitivity 150 μV/mm, (sensitivity reduced for stimulation channels (LY 4-6) for better visualization).

## Discussion

This work provides evidence that thalamocortical network neuromodulation using 4-lead systems is well tolerated and is associated with reduced seizure frequency and seizure severity in a cohort of 7 individuals with poorly localized, multifocal, eloquent cortex, or regional SN. Thalamocortical network neuromodulation is motivated by the hypothesis that combined stimulation of thalamic and cortical SN nodes may desynchronize SNs to reduce seizure burden and over time reorganize pathological networks.

Six subjects completed BTS stimulation trials (previously published as CSS) during iEEG monitoring, which allowed for optimization of electrode contact targets and stimulation parameters for subsequent chronic device implantation. Stimulation trials during iEEG provide a unique opportunity to screen and optimize stimulation with the benefit of high quality recording from up to 256 channels covering the putative SN. Trial stimulation was guided by IEDs, and seizures (all subjects had multiple seizures daily at pre-stimulation baseline). Trial stimulation produced a numerical reduction in IED rate on an individual level in 5 subjects and a non-significant trend towards reduction on a group level (median 29% reduction in hourly IED rate, range 0-87, P=0.062). All trial stimulation subjects had 50% or more reduction in seizure frequency with optimized settings.

All seven subjects were responders (≥50% reduction in seizure frequency) to chronic thalamocortical network neuromodulation, and four subjects had ≥90% seizure reduction (Figure 2). Additionally, seizure severity and life satisfaction improved for all 6 survey respondents. Quality of sleep numerically improved in 5 of 6 subjects (Figure 2), however, group level results were not significant, and statistical power was limited by the sample size. There was no clinical worsening of baseline mood. Of note, after an initial period of stimulation, subject 1 has enjoyed 4-years of sustained seizure freedom off of stimulation (inadvertently discontinued), suggesting plasticity and durable SN reorganization by chronic neuromodulation (mechanisms unknown).

Thalamocortical network neuromodulation can be performed with closed-loop, open-loop, or other novel stimulation paradigms. Current commercially available open-loop systems (off-label use) allow for 4-lead stimulation (as used here with 1 or 2 thalamus leads and 3 or 2 cortical leads). The closed-loop RNS 2-lead system allows for 1 thalamus and 1 cortical target [25], [24], [23]. There are also differences in electrode contact length, spacing, number, and total span between systems. Other factors may also play a role in choosing a stimulation paradigm, such as ability to acquire chronic brain recordings, subject preference for location of pulse generator, forthcoming rechargeable pulse generators, pulse generator replacement considerations, and local programming support.

In our cohort, 6 patients had seizure onset localized to a broad area that was classified as “regional,” which may represent distributed network dysfunction. Cortical lead-to-lead stimulation was used in 3 patients (Table 2). Lead-to-lead stimulation has been previously described[41], and allows for current to be concentrated across a regional territory bracketed or traversed by electrodes. Here, lead-to-lead stimulation was performed with interleaved settings so that each contact received leading phase cathodal stimulation. Lead-to-lead stimulation is possible with the Medtronic Intellis but not the Boston Scientific R32 IPG, while monopolar stimulation is possible with the R32 but not the Intellis IPG, which may influence system selection.

Additionally, it is unclear how open-loop “suppressive” stimulation compares with closed-loop “responsive” stimulation. Studies with RNS have established that the number of stimulations delivered far exceeds the seizure rate, which suggests an important chronic neuromodulatory effect. Interestingly, long-term analysis of RNS data has also shown that seizure suppression is positively correlated with IED suppression in individuals that respond well to RNS [42], which may further support the use of IEDs as a BTS biomarker. The relative impact of different stimulation paradigms on seizure control SN plasticity is unknown. Prior work with open-loop cortical stimulation[33] and DBS [3] support the efficacy of chronic open-loop suppressive stimulation.

Here, stimulation parameters were determined by clinical practice. There was heterogeneity in thalamic stimulation settings, including low (<10 Hz)[43], moderate (40-70 Hz) [44], and high (>100 Hz) [4]frequency stimulation (high frequency stimulation delivered on a duty-cycle) [3], all of which have had demonstrated efficacy in prior studies. The epilepsy neuromodulation stimulation parameter space is enormous and largely unexplored, and new data driven methods, such as BTS stimulation trials using short latency biomarkers are needed to efficiently screen and optimize personalized parameters sets. Future BTS applications may benefit from automated protocols, such as Bayesian optimization as demonstrated in preclinical work[45].

During chronic thalamocortical network neuromodulation, stimulation settings were individualized based on patient feedback and seizure diaries. There was transient worsening in baseline seizure frequency in two subjects. In subject 3, this was managed by reducing cortical stimulation amplitude. In subject 7 cortical stimulation was turned off at last visit with continued thalamic stimulation. Thalamocortical network neuromodulation provides the flexibility to selectively prioritize cortical or thalamic stimulation without the need for revision of the implant. Side effects of paresthesia, and acute facial twitches were reported by subject 6 and subject 7, respectively, during parameter adjustment and resolved with programming changes. The availability of segmented leads with open-loop systems allows for radially directed current steering to limit side effects in subcortical structures (of particular importance for CM stimulation) with neighboring side effect prone eloquent structures (nucleus Ventralis caudalis). One subject required pulse generator pocket revision due to delayed infection; larger studies are needed to evaluate hemorrhage and infection rates compared to 2-lead systems (3-5% and 4-9%, respectively), which we would expect to be comparable [46].

This retrospective review is limited by the nature of this study type, which lacks randomization, blinding, and a control cohort. Intracranial EEG recordings from channels undergoing BTS trial stimulation were partially obscured by stimulation artifact, and so non-stimulated neighboring contacts were used for automated IED quantification—this may underestimate the true impact of trial stimulation on SN IEDs. Future applications of BTS trial stimulation would benefit from stimulation artifact rejection techniques to preserve cerebral signals from stimulated SN channels. Change in antiseizure medication regimen between baseline and BTS trial stimulation periods in 3 subjects may impact the interpretability of our trial stimulation results for these subjects, however, findings were similar for the remaining subjects without medication change. There was considerable heterogeneity in patient factors, seizure localization, and final stimulation settings, and more work is needed to assess generalizability of these results across different SNs. The extent to which iEEG trial stimulation contributes to long-term efficacy is unclear and requires further long-term study. The off-label use of 4-port IPGs with four intracranially placed leads used here does not have established MRI safety conditions, which may impact access to future MRI imaging. Seizure severity and life satisfaction scores used a pragmatic 10 point scale to limit participant survey fatigue, instead of more comprehensive batteries such as the QOLIE-31. The 7 subject cohort is relatively small, however, this compares favorably with prior studies of thalamocortical RNS.

## Conclusion

Results from our retrospective clinical series show that thalamocortical network neuromodulation with 4-lead open-loop stimulation is a feasible, safe, and efficacious treatment for selected patients with poorly localized, regional, or multifocal SNs. Thalamocortical neuromodulation was associated with a significant reduction in seizure frequency and severity, and improved life satisfaction. BTS trial stimulation delivered during iEEG provides a unique opportunity to individualize stimulation to refine SN engagement and optimize parameters, with the benefit of high quality large channel count recordings. Future prospective controlled studies are needed to further evaluate the efficacy and generalizability of thalamocortical network neuromodulation.

## Data Availability

All data produced in the present study are available upon reasonable request to the authors

## Author Contributions

S.A.: writing – original draft; formal analysis; investigation; writing – review and editing. J.L.A.Z.: writing – review and editing, investigation. B.L. conceptualization, writing – review and editing, G.W. conceptualization, writing – review and editing. N.G. conceptualization, formal analysis, investigation, writing – review and editing. G.O., K.S., B.B., D.S., K.L., M.S., K.M., J.V.G. all made substantial contributions to writing – review and editing.

## Conflicts of interest

G.A.W., B.N.L., J.J.V.V., M.S., and B.H.B. declare intellectual property licensed to Cadence Neuroscience (BNL waived contractual rights). B.N.L. declares intellectual property licensed to Seer Medical (contractual rights waived). G.A.W. licensed intellectual property and serves on the scientific advisor board of NeuroOne, Inc. B.N.L., G.A.W, N.M.G. are investigators for the Medtronic Deep Brain Stimulation Therapy for Epilepsy Post-Approval Study. B.N.L is an investigator for the Neuroelectrics tDCS for Patients with Epilepsy Study. J.J.V.V., G.A.W., B.N.L., N.M.G. are investigators for the NeuroPace RNS NAUTILUS study. K.L. is Chief Executive Officer and co-founder, and D.S. is Chief Scientific Officer and co-founder of Cadence Neuroscience. N.M.G has consulted for NeuroOne, Inc. (funds to Mayo Clinic). B.N.L has consulted for Epiminder, Medtronic, Neuropace, and Philips Neuro (all funds to Mayo Clinic). The remaining authors declare no competing interests.

This work was approved by the Mayo Clinic Institutional Review Board. Data will be made available upon reasonable request.

## REFERENCES

1. Kalilani, L., et al., The epidemiology of drug-resistant epilepsy: A systematic review and meta-analysis. Epilepsia, 2018. 59(12): p. 2179–2193.

2. Kwan, P., et al., Definition of drug resistant epilepsy: consensus proposal by the ad hoc Task Force of the ILAE Commission on Therapeutic Strategies. Epilepsia, 2010. 51(6): p. 1069–77.

3. Morrell, M.J. and R.N.S.S.i.E.S. Group, Responsive cortical stimulation for the treatment of medically intractable partial epilepsy. Neurology, 2011. 77(13): p. 1295–304.

4. Fisher, R., et al., Electrical stimulation of the anterior nucleus of thalamus for treatment of refractory epilepsy. Epilepsia, 2010. 51(5): p. 899–908.

5. Fisher, R.S., Deep brain stimulation for epilepsy. Handb Clin Neurol, 2013. 116: p. 217–34.

6. Salanova, V., et al., The SANTE study at 10 years of follow-up: Effectiveness, safety, and sudden unexpected death in epilepsy. Epilepsia, 2021. 62(6): p. 1306–1317.

7. Peltola, J., et al., Deep Brain Stimulation of the Anterior Nucleus of the Thalamus in Drug-Resistant Epilepsy in the MORE Multicenter Patient Registry. Neurology, 2023. 100(18): p. e1852–e1865.

8. Kerezoudis, P., et al., Chronic subthreshold cortical stimulation for adult drug-resistant focal epilepsy: safety, feasibility, and technique. J Neurosurg, 2018. 129(2): p. 533–543.

9. Lundstrom, B.N., et al., Chronic subthreshold cortical stimulation and stimulation-related EEG biomarkers for focal epilepsy. Brain Commun, 2019. 1(1): p. fcz010.

10. Lundstrom, B.N., et al., Chronic Subthreshold Cortical Stimulation to Treat Focal Epilepsy. JAMA Neurol, 2016. 73(11): p. 1370–1372.

11. Lundstrom, B.N., et al., Chronic subthreshold cortical stimulation: a therapeutic and potentially restorative therapy for focal epilepsy. Expert Rev Neurother, 2017. 17(7): p. 661–666.

12. Wyckhuys, T., et al., Suppression of hippocampal epileptic seizures in the kainate rat by Poisson distributed stimulation. Epilepsia, 2010. 51(11): p. 2297–304.

13. He, X., et al., Disrupted basal ganglia-thalamocortical loops in focal to bilateral tonic-clonic seizures. Brain, 2020. 143(1): p. 175–190.

14. van Diessen, E., et al., Functional and structural brain networks in epilepsy: what have we learned? Epilepsia, 2013. 54(11): p. 1855–65.

15. Freestone, D.R., et al., The thalamocortical circuit and the generation of epileptic spikes in rat models of focal epilepsy. Annu Int Conf IEEE Eng Med Biol Soc, 2009. 2009: p. 1533–6.

16. Guye, M., et al., The role of corticothalamic coupling in human temporal lobe epilepsy. Brain, 2006. 129(Pt 7): p. 1917–28.

17. Huguenard, J.R., Prince, D.A, Basic mechanisms of epileptic discharges in the thalamus, in The Thalamus: Experimental and Clinical Aspects, M. Steriade, Jones, E.G., McCormick, D., Editor. 1997, Elsevier. p. pp. 295–330.

18. Jones, E.G., The Thalamus. 2012: Springer Science and Business Media. 1115–1175.

19. Kim, S.H., et al., Thalamo-cortical network underlying deep brain stimulation of centromedian thalamic nuclei in intractable epilepsy: a multimodal imaging analysis. Neuropsychiatr Dis Treat, 2017. 13: p. 2607–2619.

20. Martin-Lopez, D., et al., The Role of Thalamus Versus Cortex in Epilepsy: Evidence from Human Ictal Centromedian Recordings in Patients Assessed for Deep Brain Stimulation. Int J Neural Syst, 2017. 27(7): p. 1750010.

21. Paz, J.T., et al., Closed-loop optogenetic control of thalamus as a tool for interrupting seizures after cortical injury. Nat Neurosci, 2013. 16(1): p. 64–70.

22. Piper, R.J., et al., Towards network-guided neuromodulation for epilepsy. Brain, 2022. 145(10): p. 3347–3362.

23. Elder, C., et al., Responsive neurostimulation targeting the anterior nucleus of the thalamus in 3 patients with treatment-resistant multifocal epilepsy. Epilepsia Open, 2019. 4(1): p. 187–192.

24. Burdette, D., et al., Brain-responsive corticothalamic stimulation in the pulvinar nucleus for the treatment of regional neocortical epilepsy: A case series. Epilepsia Open, 2021. 6(3): p. 611–617.

25. Burdette, D.E., et al., Brain-responsive corticothalamic stimulation in the centromedian nucleus for the treatment of regional neocortical epilepsy. Epilepsy Behav, 2020. 112: p. 107354.

26. Alcala-Zermeno, J.L., et al., Centromedian thalamic nucleus with or without anterior thalamic nucleus deep brain stimulation for epilepsy in children and adults: A retrospective case series. Seizure, 2021. 84: p. 101–107.

27. Ilyas, A., et al., The centromedian nucleus: Anatomy, physiology, and clinical implications. J Clin Neurosci, 2019. 63: p. 1–7.

28. Dalic, L.J., et al., DBS of thalamic centromedian nucleus for Lennox-Gastaut syndrome (ESTEL trial). Ann Neurol, 2021.

29. Filipescu, C., et al., The effect of medial pulvinar stimulation on temporal lobe seizures. Epilepsia, 2019. 60(4): p. e25–e30.

30. Toprani, S. and D.M. Durand, Long-lasting hyperpolarization underlies seizure reduction by low frequency deep brain electrical stimulation. J Physiol, 2013. 591(22): p. 5765–90.

31. Yu, T., et al., High-frequency stimulation of anterior nucleus of thalamus desynchronizes epileptic network in humans. Brain, 2018. 141(9): p. 2631–2643.

32. Keller, C.J., et al., Induction and Quantification of Excitability Changes in Human Cortical Networks. J Neurosci, 2018. 38(23): p. 5384–5398.

33. Alcala-Zermeno, J.L., et al., Invasive neuromodulation for epilepsy: Comparison of multiple approaches from a single center. Epilepsy Behav, 2022. 137(Pt A): p. 108951.

34. Barkmeier, D.T., et al., High inter-reviewer variability of spike detection on intracranial EEG addressed by an automated multi-channel algorithm. Clin Neurophysiol, 2012. 123(6): p. 1088–95.

35. Grewal, S.S., et al., Fast gray matter acquisition T1 inversion recovery MRI to delineate the mammillothalamic tract for preoperative direct targeting of the anterior nucleus of the thalamus for deep brain stimulation in epilepsy. Neurosurg Focus, 2018. 45(2): p. E6.

36. Krauth, A., et al., A mean three-dimensional atlas of the human thalamus: generation from multiple histological data. Neuroimage, 2010. 49(3): p. 2053–62.

37. Agashe, S., et al., Centromedian Nucleus of the Thalamus Deep Brain Stimulation for Genetic Generalized Epilepsy: A Case Report and Review of Literature. Front Hum Neurosci, 2022. 16: p. 858413.

38. Rusheen, A.E., et al., Preliminary Experience with a Four-Lead Implantable Pulse Generator for Deep Brain Stimulation. Stereotact Funct Neurosurg, 2023. 101(4): p. 254–264.

39. Horn, A., et al., Lead-DBS v2: Towards a comprehensive pipeline for deep brain stimulation imaging. Neuroimage, 2019. 184: p. 293–316.

40. Alcala-Zermeno, J.L., et al., Cortical and thalamic electrode implant followed by temporary continuous subthreshold stimulation yields long-term seizure freedom: A case report. Epilepsy Behav Rep, 2020. 14: p. 100390.

41. Ma, B.B., et al., Responsive neurostimulation for regional neocortical epilepsy. Epilepsia, 2020. 61(1): p. 96–106.

42. Arcot Desai, S., T.K. Tcheng, and M.J. Morrell, Quantitative electrocorticographic biomarkers of clinical outcomes in mesial temporal lobe epileptic patients treated with the RNS(R) system. Clin Neurophysiol, 2019. 130(8): p. 1364–1374.

43. Koubeissi, M.Z., et al., Low-frequency stimulation of a fiber tract in bilateral temporal lobe epilepsy. Epilepsy Behav, 2022. 130: p. 108667.

44. Valentin, A., et al., Deep brain stimulation of the centromedian thalamic nucleus for the treatment of generalized and frontal epilepsies. Epilepsia, 2013. 54(10): p. 1823–33.

45. Stieve, B.J., et al., Optimization of closed-loop electrical stimulation enables robust cerebellar-directed seizure control. Brain, 2023. 146(1): p. 91–108.

46. Simpson, H.D., et al., Practical considerations in epilepsy neurostimulation. Epilepsia, 2022. 63(10): p. 2445–2460.

