## Supplemental Information for "Thalamocortical network neuromodulation for epilepsy"

**Hardware specifications:**

Four contact depth electrodes Medtronic 3387 (1.5 mm contact length with 1.5 mm inter-contact spacing) or an eight contact Boston Scientific Vercise DB-2202-45 segmented directional lead (1.5 mm contact length and 0.5 mm inter-contact spacing, with a total of 4 levels with the middle two levels separated into three radial segments) were implanted unilaterally into the thalamus. Four contact Medtronic 3387 or four contact Medtronic 3391 (3391 used under an IRB protocol specifically addressing the humanitarian device exemption designation; 3 mm contact length and 4 mm inter-contact spacing) or in-line eight contact Boston Scientific Vercise DB-2201-45 (1.5 mm contact length and 0.5 mm inter-contact spacing) were used to target the cortical SN regions.

| Subj. | Baseline seizure rate (per 24 hr.) | BTS stimulation trial seizure rate (per 24 hr.) | Seizure rate reduction (%) | Baseline IED rate (per hr.) | BTS stimulation trial IED rate (per hr.) | IED rate reduction (%) |
| --- | --- | --- | --- | --- | --- | --- |
| 1 | 6 | 0* | 100 | 540 | 180 | 67 |
| 2 | 8 | 0 | 100 | 634 | 636 | 0 |
| 3 | 44 | 3 | 93 | 502 | 376 | 25 |
| 4 | 2 | 0 | 100 | 2037 | 1374 | 33 |
| 5 | 25 | 11 | 56 | 1463 | 188 | 87 |
| 6 | 20 | 0 | 100 | 701 | 550 | 22 |

**Supplemental Table 1. Biomarker targeted stimulation trial results.** Baseline seizure rate is the number of seizures that occurred in the 24 hours preceding initiation of the stimulation trial. BTS stimulation trial seizure rate reflects the number of seizures that occurred during 24 hours of stimulation at optimized settings. Hourly IED rate was determined using an open-source automated detector. *Subject 1 underwent a 21-hour stimulation trial.

**Summary of biomarker targeted stimulation trial clinical data:**

Subjects 3, 5, and 6 were on stable medications throughout the baseline and trial stimulation periods.

Subject 1 received 1 mg intravenous Ativan IV at 23:30 the day prior to trial stimulation initiation, and 1 mg intravenous Ativan at 02:18 the day of trial stimulation initiation. Trial stimulation started 9 hours and 25 minutes after the second dose of Ativan, and continued for 21 hours on stable medications using a single stimulation parameter set. Stimulation was delivered through externalized chronic leads, having previously completed stereo EEG monitoring without undergoing a stimulation trial. The clinical team interpreted the stimulation trial effective as the patient “continued with the same configuration throughout the day without presenting any seizure activity or interictal activity.” Subject 1 proceeded to internalization of leads and placement of the implantable pulse generator the following day.

Subject 2 received Oxcarbazepine 600 mg twice daily at baseline, which was increased to 900 mg twice daily during the 42 hour trial stimulation period. Two stimulation contact configurations and stimulation amplitudes were trialed.

Subject 3 remained on home medications throughout the baseline and trial stimulation periods. Subject 3 completed 44 hours of trial stimulation, during which two stimulation contact configurations, and both continuous and duty cycle stimulation was trialed.

Subject 4 received an intravenous load of lacosamide prior to stimulation functional mapping, which was 5 hours before initiation of trial stimulation. Antiseizure medications were otherwise stable between baseline and trial stimulation periods. Subject 4 was seizure free during 29 hours of trial stimulation. Three stimulation configurations were used.

Subject 5 remained on home medications throughout the baseline and trial stimulation periods. Subject 5 underwent 48 hours of trial stimulation, with two stimulation contact variations, two stimulation pulse frequency variations, and multiple stimulation amplitude adjustments.

Subject 6 remained on stable medications during the baseline period and 51 hours of trial stimulation. Subject 6 had zero seizures during the stimulation trial. Three stimulation contact variations were used, and stimulation amplitudes underwent multiple adjustments.
